# Heavy-tailed sexual contact networks and the epidemiology of monkeypox outbreak in non-endemic regions, May 2022

**DOI:** 10.1101/2022.06.13.22276353

**Authors:** Akira Endo, Hiroaki Murayama, Sam Abbott, Ruwan Ratnayake, Carl A. B. Pearson, W. John Edmunds, Elizabeth Fearon, Sebastian Funk

## Abstract

A global outbreak of monkeypox across non-endemic regions including Europe and North America was confirmed in May 2022. The current outbreak has shown distinct epidemiological features compared with past outbreaks in non-endemic settings, most notably its observed rapid growth and predominant spread among men who have sex with men (MSM). We use a branching process transmission model fitted to empirical sexual partnership data in the UK to show that the heavy-tailed nature of the sexual partnership degree distribution, where a small fraction of individuals have disproportionately large numbers of partners, can explain the sustained growth of monkeypox cases among the MSM population despite the absence of such patterns of spread in past outbreaks. We also suggest that the basic reproduction number (*R*_0_) for monkeypox over the MSM sexual contact network may be substantially greater than 1 for a plausible range of assumptions, which poses a challenge to outbreak containment efforts. Ensuring ongoing support and tailored public health messaging to facilitate prevention and early detection among MSM with a large number of sexual partners is warranted.

## Main text

In May 2022, multiple countries in Europe, North America and elsewhere reported clusters of monkeypox cases (*1*–*4*). As of 31 May 2022, a total of 728 confirmed and suspected cases have been reported in over 25 countries from previously non-endemic regions (*5*). To date, the reported cases are predominantly, but not exclusively, among young males without a travel history to endemic regions in Central and West Africa (*2, 3*). The initial epidemiological investigations suggest a link with sexual contact among men who have sex with men (MSM) (*1–3, 6*). Prior to the current outbreak, monkeypox infections had been assumed to be primarily caused by exposure to animal reservoirs but human-to-human transmissions via direct routes including skin-to-skin contact, bodily fluids and respiratory droplets have also been documented (*7, 8*). Sexually-associated exposure to skin lesions, droplets and fomites could plausibly be a high risk for transmission, whether monkeypox is truly sexually transmissible (e.g. via semen) or not. Previous studies of monkeypox outbreaks indicate around 10% secondary attack risk (SAR) among household members without smallpox vaccination (*7*–*9*); the smallpox vaccine has been shown to be protective against monkeypox with estimated effectiveness of 85% (*9*). Investigation of previous outbreaks in Central and West Africa identified a relatively limited proportion of cases of human-to-human transmission, with at most seven generations observed (*7, 8, 10, 11*), and previous estimates of the basic reproduction number (*R*_0_) for monkeypox have been below 1 even in unvaccinated populations (*8, 9*). Sporadic monkeypox outbreaks associated with imported animals or imported cases have been observed in non-endemic regions (*12*–*16*) but subsequent human-to-human spread was rarely observed. Prior to the current outbreak, only one healthcare worker and two household contacts of an imported case had been identified as likely secondary cases in non-endemic settings (*14, 16*).

The current spread of monkeypox in non-endemic regions appears in stark contrast to these previous events. The vast majority of cases have no documented exposure to animals or travel history to endemic settings. The rapid growth in notified cases and geographical dispersal suggest substantial human-to-human transmission, rather than incidence driven by spillover from an animal reservoir. This is also the first widespread outbreak of monkeypox predominantly in MSM with suggested sexually-associated transmission (*1, 17*), although higher prevalence in young males and frequent observation of genital lesions have also been documented in a recent outbreak in Nigeria (*11, 18, 19*). Proposed explanations for the novel character of the current outbreak include increased importation, undetected community-wide transmission, viral evolution, and increased susceptibility due to the end of smallpox vaccination (*6, 16, 17*). While these theories are consistent with some aspects of the current observation, most of them are not strongly supported by external (if indirect) evidence nor do they provide a coherent explanation on why a similar monkeypox outbreak involving substantial human-to-human transmission in a focal, rather than generalised, population had not arisen from the series of importation events documented in non-endemic settings starting in 2003 (*12*–*16*).

Here we show that transmission over a sexual contact network empirically characterised by a heavy-tailed partnership distribution can reasonably explain the rapid growth of human-to-human transmissions in the current monkeypox outbreak despite the absence of such patterns of spread in the past. Specifically, it is plausible that monkeypox has had a substantial transmission potential in the MSM sexual contact network but that due to the small cumulative number of imported cases in non-endemic settings, it had not reached high degree members of this network from whom onwards transmission was most probable.

Previous work on sexual partnership distributions (i.e. degree distribution of sexual contact networks) often fitted Pareto distributions to the reported number of partners over a specific time window (e.g. over a year) (*20, 21*) However, Pareto distributions exhibit the scale-free property, which causes the modelled networks to have some individuals with impossibly high numbers of partners and *R*_0_ defined in the networks to tend to infinity (*22*). We found that Pareto distributions do not describe existing datasets of MSM partnerships well (see *Supplementary materials*). The Weibull distribution, suggested as an alternative distribution that also has a heavy-tailed shape (*20*), does not exhibit the unphysical features of the Pareto distribution. Using Weibull distributions fitted to the empirical data on same-sex and opposite-sex sexual partnerships of the UK population aged 18–44 (from the National Surveys of Sexual Attitudes and Lifestyles; Natsal (*23*–*25*)), we constructed a branching process model of transmission over sexual contact networks. Following (*22*), we assumed that individuals can become infected at a probability proportional to their network degree (i.e. those with a large number of partners are more likely to be chosen. We also assumed an infectious period of 21 days for monkeypox based on the documented duration of illness (*26– 28*), which we also varied in our sensitivity analysis in *Supplementary materials* to account for variation and possible behaviour changes in symptomatic individuals.

Using this model, we simulated sexually-associated outbreaks of monkeypox in MSM and non-MSM populations, under varying assumptions for the risk of transmission between sexual partners (SAR per sexually-associated contact during the infectious period) between 0–100% in the absence of empirical data on this parameter. Starting from a specified number of initial cases, we simulated the number of cases in each generation of transmission over MSM and non-MSM sexual contact networks. For the non-MSM sexual network, we assumed that the initial cases have equal chances of being male or female and that subsequent generations of infection alternate between heterosexual (HS) men and women. Women who have sex with women were not considered in our analysis as their partnership distribution suggested a substantially lower transmission potential than the HS network. In the model, we considered only sexually-associated transmission over separate sexual contact networks of MSM and non-MSM and did not explicitly model other transmission routes (non-sexually-associated skin-to-skin contact, respiratory droplets, fomites, etc.) or links between MSM and non-MSM sexual contact networks, except for the initial cases. We discuss transmission dynamics of monkeypox as a mixture of these transmission routes in a separate analysis using the next generation matrix (*29*). The methodology used is described in more detail in the *Supplementary materials*.

We first simulated the probability of observing a chain of transmission of a size equal to or greater than the current global monkeypox outbreak (728 cases as of 31 May 2022) in the MSM population generated from a given number of initial cases. We considered three scenarios for the profile of introduction events: (1) The initial cases in the MSM population acquired infection via a sexually-associated route, i.e. the numbers of their sexual partners are preferentially drawn from the tail of the sexual partnership distribution; (2) The initial cases in the MSM population were non-sexually-associated and therefore their partnership degrees were drawn from across the full distribution; (3) Initial cases were from the general population (of which 2% were assumed to be sexually-active MSM based on the Natsal datasets) who acquired infection via non-sexually-associated routes. We then simulated the probability of the current outbreak leading to a major outbreak over the MSM sexual contact network. For comparison, outbreaks over the non-MSM sexual contact network given specified numbers of initial cases (either sexually-associated or non-sexually-associated) were also simulated. Our estimates suggest that with a range of sexually-associated SAR values comparable to or greater than the previous estimates of household SAR (*7*–*9*), even one event of sexually-associated transmission to the MSM population (scenario 1) is consistent with a high likelihood (20–50%) of observing an outbreak of the current size or greater (Fig. 1A). The likelihood becomes smaller (although not negligible) if non-sexually-associated exposure (e.g. exposure to animals or non-sexual direct contact with cases; scenario 2) is involved in the introduction to the MSM population (Fig. 1B). On the other hand, 50–100 or more non-sexually-associated initial cases from the general population (scenario 3) would have been necessary for the likelihood of an outbreak of the current size in the MSM population to be around the order of 1–20% (Fig. 1C).

**Fig. 1.**
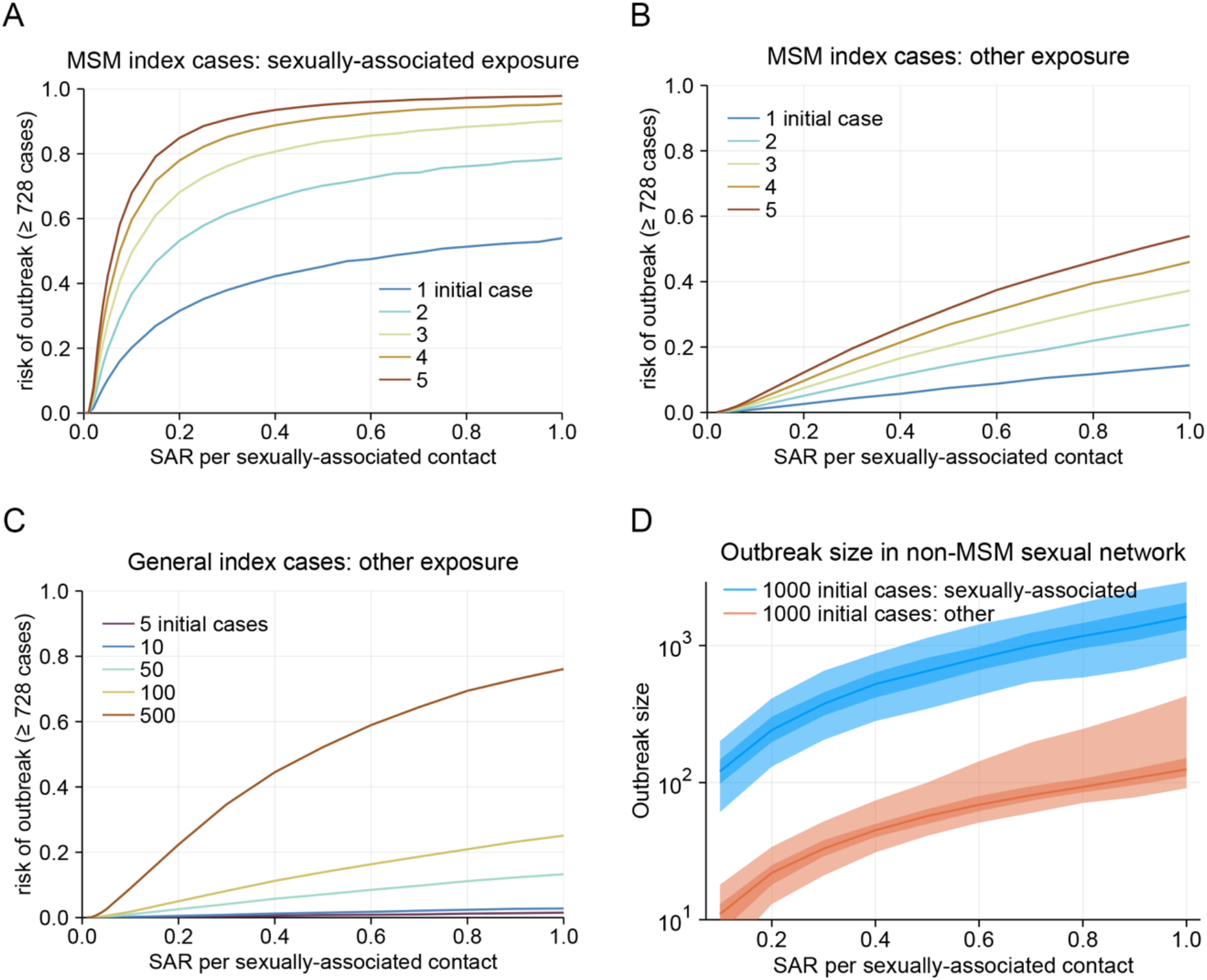
Likelihood of observing an outbreak given introduction events of different profiles. (A–C) The likelihood of observing an outbreak of the current size (728 cases) or greater over the MSM sexual contact network given initial cases who are: (A) MSM with sexually-associated exposure; (B) MSM with non-sexually-associated exposure; (C) random cases from the general population with non-sexually-associated exposure. The likelihood was computed from 100,000 simulations for each value of SAR varied between 0% and 100%. (D) Simulated outbreak sizes over non-MSM sexual contact network given 1,000 non-MSM initial cases who have had sexually-associated exposure (blue) or non-sexually-associated exposure (red). Mid-lines and thick and thin ribbons represent median, interquartile and 95% quantile ranges, respectively.

These results suggest that a very small number of sexually-associated transmissions among the MSM population are sufficient to cause a large outbreak over the MSM sexual network— as we now appear to observe—but that the number of non-sexually-associated imported cases required for the virus to achieve the first few instances of sexually-associated transmission among MSM is relatively large. The cumulative number of documented imported cases in non-endemic settings had been up to around 100 prior to May 2022 (*12*–*16*); it is therefore unsurprising that introduction to the MSM population in non-endemic settings has never been observed previously, assuming that the importations had been mostly non-sexually-associated cases from the general population. The current outbreak in the MSM population may have been caused by an eventual introduction after following non-sexually-associated importations or, alternatively, by one or more sexually-associated importations acquired in the endemic setting. In the latter case, a sexually-associated outbreak among MSM might also be ongoing in the endemic settings, which warrants further surveillance. All scenarios projected that, without interventions or changes to sexual behaviour, a major outbreak in the MSM population (defined as ≥ 10,000 cases excluding initial cases) is highly likely given the current outbreak size (Table 1). In contrast, sustained transmission in the non-MSM population is unlikely in all scenarios considered (Table 1) due to the less heavy tail of the corresponding partnership distribution, although from 10 up to 10,000 additional cases may be observed if a substantial number of infections are introduced into the non-MSM sexual contact network (Fig. 1D).

**Table 1.** Likelihood of an outbreak over MSM or non-MSM sexual contact network given different numbers and profiles of introduction events.

| SAR | Likelihood of 728+ cases in MSM given 1 introduction event |  |  | Likelihood of 10000+ cases given multiple introductions |  |
| --- | --- | --- | --- | --- | --- |
|  | S-A event (MSM) | Non-S-A event (MSM) | Non-S-A event (Gen. pop.) | 728 S-A events in MSM | 1000 non-S-A events in non-MSM |
| 5% | 10% | 0.25% | 0.003% | 100% | < 0.001% |
| 10% | 20% | 0.9% | 0.02% | 100% | < 0.001% |
| 20% | 31% | 2.6% | 0.04% | 100% | < 0.001% |
| 50% | 45% | 7.3% | 0.16% | 100% | < 0.001% |

SAR: secondary attack risk; MSM: men who have sex with men; S-A: Sexually-associated; Gen. pop.: general population.

The projected values of *R*_0_ were almost always above 1 in the MSM sexual network for a range of sexually-associated SAR, while *R*_0_ for the non-MSM sexual network was found to be below 1 unless SAR was nearly 100% (Fig. 2A). The potentially high *R*_0_ for the MSM sexual network is particularly concerning because it can pose challenges to the control efforts (Fig. 2B). Contact tracing and ring vaccination approaches may need to identify almost all contacts of a case to bring the epidemic under control (which would not be easily achievable in practice (*2*)) because untraced transmission may well lead to other sustained transmission chains. Extensive contact tracing is now being conducted in many places with cases, however since most of the cases included in our analysis probably occurred before contract tracing was reinforced, we cannot yet estimate the effectiveness of these ongoing efforts. Another possible approach would be to focus resources on identifying acceptable and effective means of preventing transmission among those men with the highest number of sexual partners, which could have a disproportionate effect on transmission overall. The *R*_0_ may sharply decrease if control efforts are effective in the tail part of the partnership distribution (represented by the upper 1^st^ percentile among those with at least one partner over the infectious period) (Fig. 2C). This would also lower the required intensity of other measures to achieve outbreak control (Fig. 2D). Substantial impact may be achievable by preventing instances in which a single initial case causes more than 10 secondary cases, where the contact patterns of the majority of the individuals are minimally affected (Fig. 2E).

**Fig. 2.**
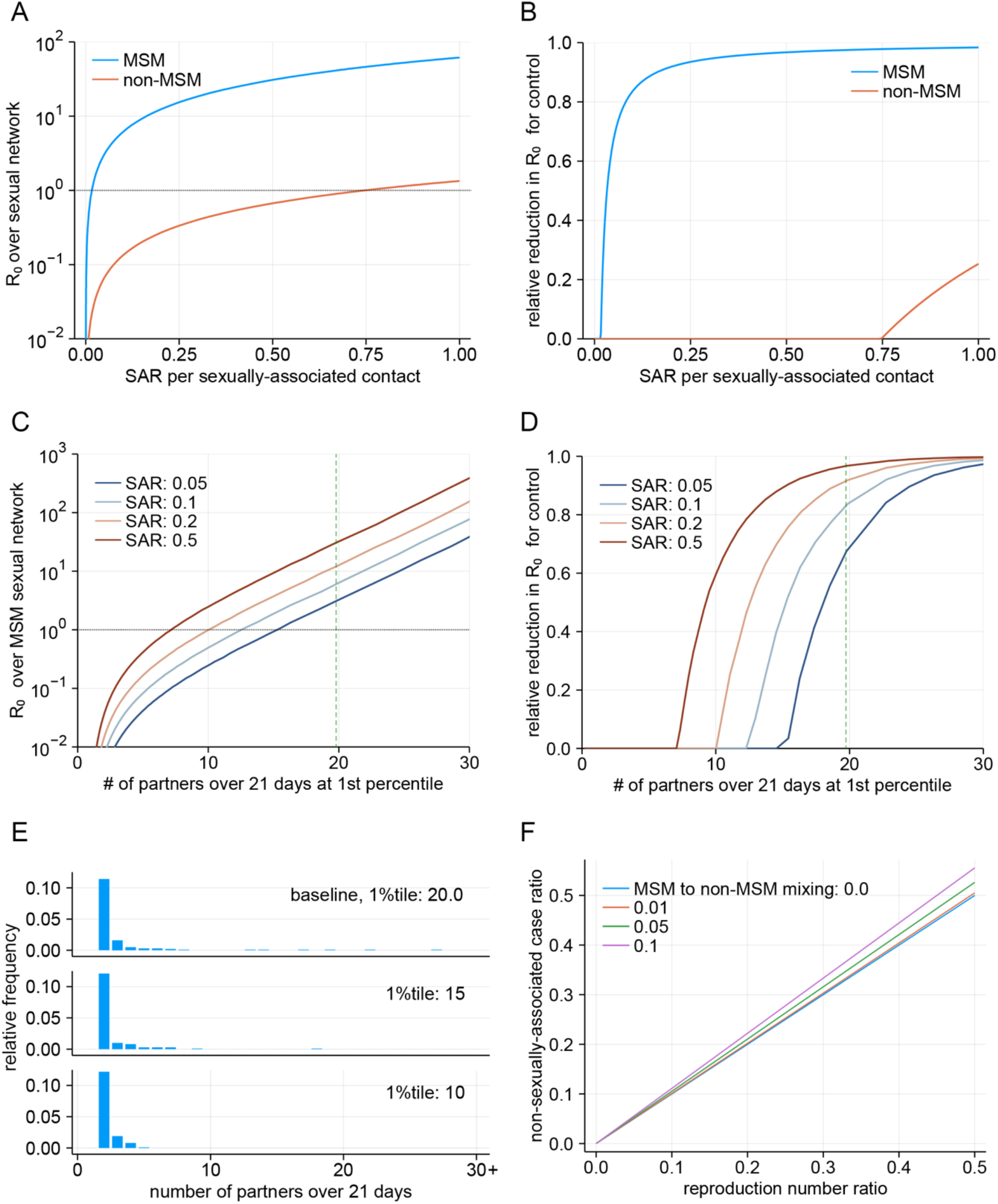
Basic reproduction number (*R*_0_) of monkeypox over sexual contact networks and control. (A) Projected *R*_0_ over the MSM and non-MSM sexual contact networks based on the Natsal sexual partnership datasets. The dotted horizontal lines denote the epidemic threshold (*R*_0_ = 1). (B) Relative reduction in *R*_0_ required to bring the outbreak under control. (C) Projected *R*_0_ over the MSM sexual contact networks with different levels of the partnership distribution tail. Holding the body shape of the distribution (see *Supplementary materials* for details) constant, the upper 1^st^ percentile of the number of 21-day sexual partners (among MSM with at least one partner over 21 days) was varied. Dashed green lines indicate the upper 1^st^ percentile of the Weibull distribution fitted to the Natsal datasets (baseline). (D) Relative reduction in *R*_0_ required for control with different levels of the partnership distribution tail. (E) Simulated 21-day sexual partnership distributions among MSM with different levels of partnership distribution tail. The original distribution fitted to the Natsal datasets (upper 1^st^ percentile: 20.0) is shown as a baseline. (F) The relationship between the reproduction number ratio and the asymptotic proportion of non-sexually-associated cases among total cases. The reproduction number ratio is defined as the ratio between the reproduction number via non-sexually-associated routes of transmission and that of the sexually-associated route. The “MSM to non-MSM mixing” parameter denotes the average proportion of non-MSM sexual partners a typical sexually-associated MSM case would have.

The analysis presented here only considered a single outbreak over either the MSM or non-MSM sexual contact network with a given number of introductions. However, understanding the disease dynamics as a mixture of interacting populations via multiple modes of transmission is crucial in projecting possible future scenarios, especially given the known or suggested non-sexually-associated routes of transmission including via droplets, fomites or aerosols (*8, 30*). One of the key questions is whether the current monkeypox outbreak can be sustained in the general community via non-sexually-associated routes, i.e. whether the *R*_0_ corresponding to non-sexually-associated transmission is above or below 1. Although we are unable to answer this question because the presence of non-sexually-associated epidemiological links in the current outbreak is so far largely unknown, we propose a possible approach to inferring the role of such transmission. We show in Fig. 2F that the proportion of cases without a sexually-associated epidemiological link among total cases will approximately approach the ratio between *R*_0_’s over the MSM sexual network and general non-sexually-associated transmission routes as the outbreak progresses. One should be able to conclude that the non-sexually-associated *R*_0_ for monkeypox is substantially lower than *R*_0_ over the MSM sexual network if the proportion of non-sexually-associated cases remained low in the future; however, caution is warranted as even in that case the general transmission *R*_0_ may still be above 1 if *R*_0_ over the MSM sexual network is as high as suggested in some scenarios presented in our analysis.

Without needing a novel hypothesis, our results propose a simple but coherent explanation of the first rapidly growing sexually-associated monkeypox outbreak in non-endemic regions in history, with a suggested link to the MSM population, using empirical sexual partnership data. We also suggested that *R*_0_ over the MSM sexual network can be substantially higher than previous estimates in non-sexually-associated contexts if the sexually-associated SAR is comparable to or greater than the household SAR, highlighting an immediate need for control efforts to inform and protect the MSM community. Self-sustained transmission over the entire non-MSM sexual network or via non-sexually-associated routes appears less likely, although a substantial number of cases may still be observed if the outbreak continues to grow in the subsets of sexual contact networks at a higher risk of transmission. Control efforts such as contact tracing and ring vaccination need to achieve high effectiveness given the large *R*_0_ values we have estimated; focused public health messaging and support for individuals with a large number of sexual partners would complement these approaches to bring the outbreak under control.

Our conclusions hinge on the assumed parameters from previous outbreaks with different transmission routes, including the SAR and infectious period of monkeypox, as well as the observed characteristics of the sexual partnership distribution in the UK and accompanying assumptions. We modelled the global transmission of monkeypox over a single connected sexual contact network fitted to the datasets of the UK population aged 18–44, while populations not represented by those datasets may be more or less vulnerable to the sexually-associated monkeypox outbreak due to different partnership patterns and *R*_0_ values (which may also explain the limited observation of possible sexually-associated outbreaks in endemic countries previously, although this may be in part due to insufficient case ascertainment). However, our sensitivity analysis using only case counts within the UK yielded almost identical results (Table S3) and the same approach could also be applied to other population settings where sexual partnership data is available. We did not consider the possibility of degree assortativity or clustering, which would lead to more densely-clustered local subnetworks than we modelled. The presence of these properties could reduce the final outbreak size due to quick depletion of high-degree susceptibles (*31*). It is also plausible that there could be core parts or clusters of the non-MSM sexual contact networks over which transmission could be sustained, which is not captured by modelling transmission over the non-MSM partnership distribution as a whole. Finally, because of the limited sample size of MSM partnerships in the Natsal datasets (*N* = 409), uncertainty remains around their *R*_0_ values (see the sensitivity analysis in *Supplementary materials*). Our estimates should be viewed as a qualitative projection rather than precise estimates of *R*_0_. Future empirical evidence from the current outbreak and estimates of key epidemiological parameters as well as the effectiveness of interventions will inform our projections on the current and future epidemiology of the monkeypox outbreak.

## Data Availability

The underlying data is available from UK Data Service (https://ukdataservice.ac.uk) provided the End User Licence Agreement.

https://beta.ukdataservice.ac.uk/datacatalogue/series/series?id=2000036

## Acknowledgements

The authors thank Kosuke Yasukawa and Toshibumi Taniguchi for their expert comments, Chris I. Jarvis, Emily S. Nightingale and Nicholas G. Davies for their inputs on literature and Nicholas G. Davies and Timothy W. Russel for feedback on the early version of the manuscript. AE is supported by JSPS KAKENHI (22K17329), JSPS Overseas Research Fellowships and foundation for the Fusion Of Science and Technology. SA and SF are supported by Wellcome Trust (210758/Z/18/Z). CABP is supported by the Innovative Medicines Initiative 2 (IMI2) Joint Undertaking between European Union Horizon 2020 Research and Innovation Programme and the European Federation of Pharmaceutical Industries and Associations [EBOVAC3: grant number 800176]. RR is supported by a Doctoral Foreign Study Award from the Canadian Institutes of Health Research (award number DFS-164266). EF is supported by the Medical Research Council, UKRI (MR/S020462/1).

## Supplementary materials

### Materials and methods

#### Data source

We used the distribution of the self-reported number of sexual partners over one year in three of the UK National Survey of Sexual Attitudes and Lifestyles (Natsal) datasets: Natsal-2 (1999–2000; 12,110 participants) (*23*), Natsal-3 (2010–2012; 15,162 participants) (*24*) and Natsal-COVID (2020; 6,654 participants) (*25*). We used the number of reported partners by men and women aged 18–44 who reported at least one sexual partner (same-sex and opposite-sex) over a year in each of the datasets. We defined men and women who reported at least one same-sex partnership over one year as sexually-active men who have sex with men (MSM) and women who have sex with women (WSW), respectively. We assumed that the reported opposite-sex partners in men and women are almost representative of the sexual partnership of heterosexual (HS) individuals. To maximise the sample size for the analysis (especially for MSM, who accounted for about 4% of the male respondents), we combined multiple datasets if the model fit supported the assumption that the samples are derived from the identical distribution (see *Additional results*), which resulted in 409, 290, 7,278 and 4,623 samples used in the estimation of partnership distributions for MSM, WSW, HS men and HS women, respectively.

Of these Natsal datasets, the Natsal-COVID dataset warranted caution: sexual partnership data in the Natsal-COVID dataset was obtained from 29 July to 10 August 2020, three months into the first lockdown in the UK. While a significant minority reported change in their sexual activity during the lockdown (*32*), the questionnaire asked the number of partners “in the last year” and the relative impact of the three months under lockdown on the responses to this question is unclear (some people might also have interpreted “the last year” as the entire 2019, which was then not under the impact of lockdown). The results using the Natsal-2/3 datasets (pre-COVID data) and the Natsal-COVID dataset (COVID wave 1 data) were compared in our sensitivity analysis (*Sensitivity analysis 3*).

The outbreak size as of 31 May 2022 of the monkeypox outbreak in non-endemic regions (globally) and that in the UK was collected from the Global.health public dashboard (*5*). We aggregated both the confirmed and suspected number of cases registered in the dashboard.

#### Ethical review

This study was approved by the London School of Hygiene & Tropical Medicine ethics committee (reference number: 27985).

#### Model and simulation procedure

We modelled the branching process of monkeypox transmission over MSM and non-MSM sexual contact networks as follows. Although we do not specify the population (MSM, HS men or HS women) for notational convenience in the following formulation, the same framework was applied to both groups. Let an integer *D*^*1Y*^ be a random variable representing the degree of an individual in a sexual partnership network over a year. We assumed that the degree distribution follows a truncated Weibull distribution for the region *x* ≥ 1, i.e.:

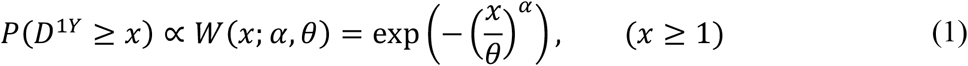

where *α* and *θ* are the shape and scale parameters of the Weibull distribution. We assumed that individuals reporting at least one partner over a year are potentially sexually active over the 21 days of infectious period of monkeypox (*26*–*28*). We estimated the parameters *α* and *θ* for same-sex/opposite-sex partnerships in men and women by fitting the Weibull distributions to the empirical degree distribution observed in Natsal datasets using Markov-chain Monte Carlo (MCMC) (see *Additional results* for details). The data was only publicly available for those with at least one sexual partner and thus *p*_0_ was not estimated, which is a nuisance parameter and does not affect the discussion hereafter. Among the three subcategories of non-MSM (HS men, HS women and WSW), HS men had the most heavy-tailed distribution, followed by HS women and WSW. This suggests that WSW, who accounted for about 3% of female respondents, have a sexual contact network with the least transmission potential and thus minimally contribute to overall transmission among non-MSM. We therefore represented the non-MSM sexual network by the partnership distributions of HS men and women. In the following simulation, we used posterior median estimates from the MCMC samples: See *Additional results* section for the estimation procedure.

The distribution of the expected number of partners over 21 days among sexually active individuals was then modelled by a continuous truncated Weibull distribution obtained with a scaling factor of 21/365, i.e.:

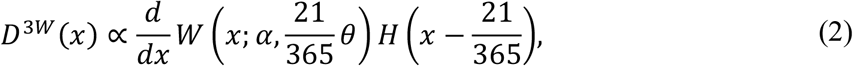

where *H*(·) is the Heaviside step function. Assuming a proportionality between the risk of infection and the number of partners, the distribution of the mean network degree over 21 days among sexually-associated monkeypox cases will follow a density:

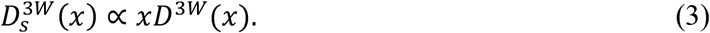

We simulated the branching process of an infection cluster originating from a given number of initial cases and estimated the probability that the final outbreak size *C*_F_ (excluding the initial cases) is equal to or greater than *x*, i.e. *P*(*C*_*F*_ ≥ *x*), by 100,000 simulations. Each case was assumed to transmit the virus to their sexual partners at a constant probability per partner (i.e. secondary attack risk; SAR) over their infectious period. The number of sexual partners of a case, *n*, was drawn from a Poisson distribution whose mean follows either *D*^3*W*^(*n*) (sexually active case with non-sexually-associated exposure), 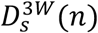(cases with sexually-associated exposure) or a zero-inflated distribution (1 − *p*_*A*_)*δ*(*n*) + *p*_*A*_*D*^3*W*^(*n*) (random case from the general population with non-sexually-associated exposure), depending on the type of the initial cases. The mixture weight *p*_*A*_ represents the proportion of sexually active individuals in the general population. We used *p*_*A*_ = 0.02 for MSM, *p*_*A*_ = 0.86 for HS men and *p*_*A*_ = 0.89 for HS women, respectively, referring to the proportion of individuals reporting at least one partner in the Natsal datasets. The number of secondary transmissions was then drawn from a binomial trial with a size *n*-1 (sexually-associated case, who was infected by one of their sexual partners) or *n* (non-sexually-associated case) with a probability equal to SAR. We iterated this transmission propagation process for each case over generations of the branching process (using a single partnership distribution for MSM throughout and alternating distributions between generations for non-MSM) until the cluster either becomes extinct or reaches the specified upper limit.

#### Reproduction number over a network

The basic reproduction number (*R*_0_) over a sexual contact network can be computed using the mean excess degree (*33*), which is given for a truncated Weibull distribution with a lower limit *l* as:

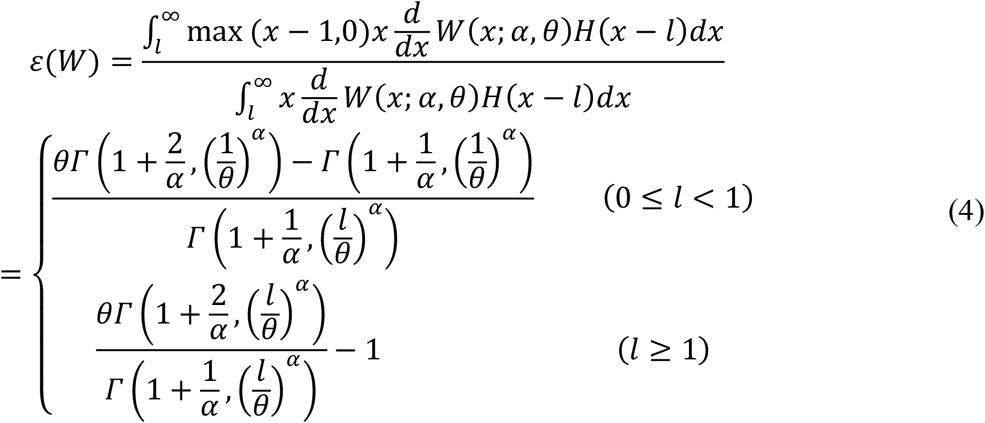

where *Γ*(*a, b*) is the incomplete gamma function. The *R*_0_’s for the MSM and non-MSM sexual contact networks are then defined as

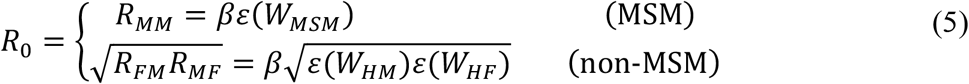

where *β* is sexually-associated SAR and *W*_MSM_, *W*_HM_, and *W*_HF_ are the Weibull distributions corresponding to MSM, HS men and HS women, respectively. We refer to *R*_FM_ and *R*_MF_ as the one-way HS reproduction numbers, which represent the mean number of secondary female cases caused by a typical male case and secondary male cases caused by a typical female case, respectively.

#### Controlling the tail of a sexual partnership distribution

A truncated Weibull distribution can be compared with a Pareto distribution, which can be represented as a linear line in a log-log plot. Let *y*_*P*_ and *y*_*W*_ be the upper cumulative density of a Pareto and Weibull distribution both defined for *x* ≥ 1. We then get

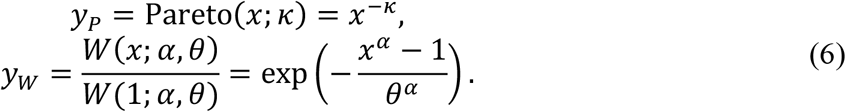

By a log-log transform where *X* = log(*x*) and *Y* = log(*y*), the above relationships can be rearranged as:

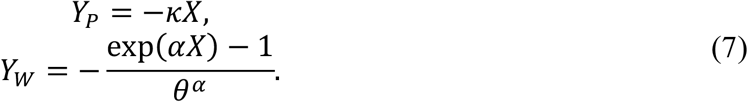

Note that for *αX* << 1 we have the following approximation for *Y*_*W*_:

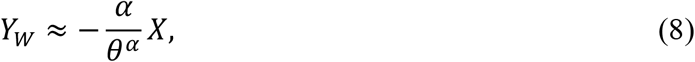

which gives a Pareto approximation for the body (i.e. non-tail part) of a truncated Weibull distribution. We assumed that the Pareto-approximation parameter for a Weibull distribution 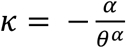 is kept constant while the tail length of the sexual partnership distribution is controlled by *α* due to interventions for individuals with a large number of partners, and then estimated the *R*_0_ reflecting different levels of control on transmission among those with the highest number of partners (represented by the upper 1^st^ percentile of those with at least one partner over the infectious period). *R*_0_ for different combinations of parameters *α* and *κ* are also explored in *Additional results*.

#### Next generation matrix for mixed modes of transmission over MSM and non-MSM networks

To account for the impact of non-sexually-associated transmission routes (e.g. non-sexual skin-to-skin contacts, droplets and fomites) on the overall transmission dynamics as well as the introduction of transmission between MSM and non-MSM sexual contact networks, we used the next generation matrix approach (*29*). Let 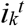 be the number of cases of type *k* in an infection generation *t*, where *k* represents MSM cases with sexually-associated exposure (*k* = 1), heterosexual male (*k* = 2) and heterosexual female (*k* = 3) cases with sexually-associated exposure and non-MSM cases (both sexes combined; *k* = 4) without sexually-associated exposure. MSM cases without sexually-associated exposure were excluded due to their relatively negligible representation among MSM cases. We represented the reproduction process between a successive pair of infection generations using a next generation matrix *K* as:

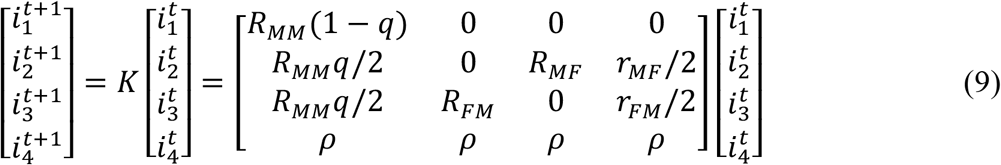

where *r*_*FM*_ and *r*_*MF*_ correspond to the mean partnership degree of a random non-MSM male and female case (i.e. acquired via non-sexually-associated exposure) and *r* is the reproduction number for general non-sexually-associated routes. The average proportion of the number of non-MSM sexual partners that a sexually-associated MSM case would have among the total partners is given as *q*, which we refer to as the MSM to non-MSM mixing ratio. On the other hand, sexually-associated transmission from non-MSM cases to the MSM population was considered negligible compared to the magnitude of sexually-associated transmission among MSM. We also assumed that the risks of transmission via sexually-associated and other routes are mutually independent such that the number of partners among non-sexually-associated non-MSM cases follows the distribution among the general population. When the generation interval is shared between sexually-associated and other routes of transmission, the relative magnitudes of 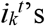 tend to the eigenvector of *K*.

All the analysis was conducted either in Julia v.1.7.2 or R v. 4.0.2. Source codes are available on a GitHub repository: (https://github.com/akira-endo/monkeypox_heavytail).

### Additional results

#### Parameter estimation for Weibull distributions for sexual partnerships from the Natsal datasets

To parameterise the branching process model of transmission among the MSM and non-MSM populations, we fitted the Weibull distribution in Equation (1), truncated at *x* = 1, to the reported number of partners in Natsal datasets (who reported at least one partner over the previous year) using MCMC. Weibull distributions are usually characterised by two parameters: shape *α* and scale *θ*; however, to preclude a high correlation between parameters in posterior samples, we used an alternative parameterisation (*α, κ*), where 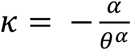 (see Equation (8))., We estimated the parameters separately for four different partnership distributions: same-sex and opposite-sex partnerships reported by men and women. For each of the partnership category, we used different subsets of the dataset that we assumed to be the most representative of the current sexual behaviours. These selections were informed by the model comparison described in the next section.

We employed a weakly-informed prior HalfNormal(*μ* = 0, *σ* = 10) except for *α* for the same-sex partnership reported by men, which had a limited sample size to fully inform this parameter (Table S1). We set a prior for this parameter based on the maximum number of partners observed in the gonococcal resistance to antimicrobials surveillance programme (GRASP) dataset: a maximum of 700 partners among 691 samples (Table S3). The likelihood for this observation assuming a truncated Weibull distribution with a fixed *κ* = 0.6 (corresponding to the reported Pareto exponent parameter in (*21*)), assigned a weight of 0.1 to prioritise the fit to the Natsal datasets, was used as a prior.

We used the Hamiltonian Monte Carlo algorithm with No-U-Turn-Sampler (via {rstan} package run in R v.4.0.2) and obtained 15,000 samples from five chains after discarding the first 2,000 samples as warm-up. The resulting MCMC samples showed an R-hat statistic of below 1.01 and an effective sample size of at least 2,000. Posterior median estimates and 95% credible intervals are shown in Table S1. The same-sex partnership reported by women had a less heavy tail (indicated by large values of *κ*) similar to the opposite-sex partnership reported by women; since transmission among WSW does not involve other populations with a heavier tail (as opposed to HS women), sustained transmission over the WSW sexual contact network is expected to be less likely than the HS population. Their estimates are shown in Table S1 for completeness but not used in our analysis. The observed and simulated distributions for MSM and HS partnerships are shown in Fig. S1.

**Table S1.** Maximum number of sexual partners among the MSM population in empirical and simulated datasets assuming a Pareto distribution.

| Category of partners | Natsal dataset used | Sample size | Shape parameter ($\alpha$) | Pareto-approximation parameter ($\kappa$) |
| --- | --- | --- | --- | --- |
| Same sex, reported by men | 2/3/COVID | 409 | 0.10 (0.02–0.19) | 0.77 (0.66–0.88) |
| Opposite sex, reported by men | 2/3 | 7,278 | 0.01 (0.00–0.04) | 1.68 (1.63–1.73) |
| Same sex, reported by women | 2/3/COVID | 380 | 0.01 (0.01–0.07) | 2.08 (1.85–2.31) |
| Opposite sex, reported by women | 3 | 4,623 | 0.01 (0.00–0.03) | 2.14 (2.07–2.21) |

Median estimates and 95% credible intervals are shown.

**Fig. S1.**
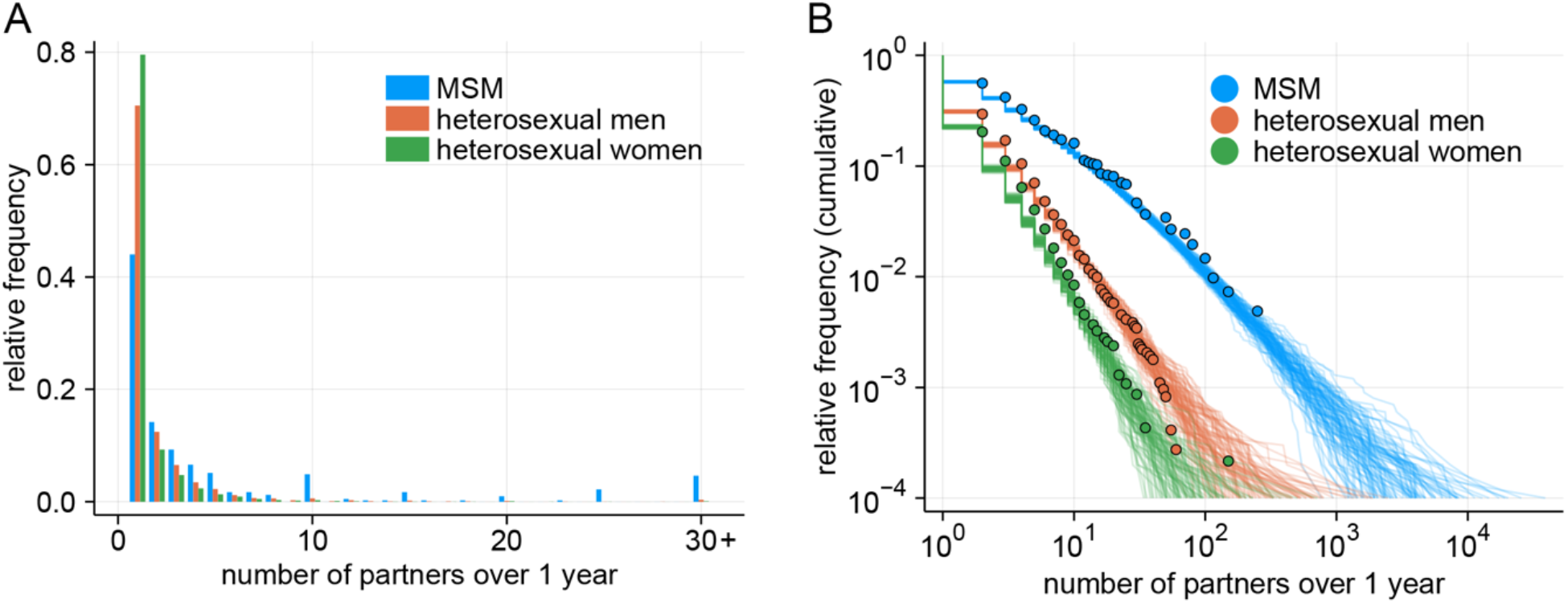
Observed and simulated partnership distributions for MSM and HS populations. (A) Observed distributions for MSM and HS sexual partnerships over 1 year in the Natsal datasets (MSM: Natsal-2/3/COVID; HS men: Natsal-2/3; HS women: Natsal-3). (B) Comparison between observed and simulated partnership distributions. For each of the populations, 100 simulated datasets (each containing 10,000 samples) were drawn from the Weibull distributions parameterised by the posterior median estimates.

#### Model comparison to check for temporal changes between Natsal datasets

In our parameter estimation, we aimed to combine multiple Natsal datasets (chosen from Natsal-2, Natsal-3 and Natsal-COVID, conducted in 1999–2000, 2010–2012 and 2020, respectively) to ensure sufficient sample sizes (in particular for MSM), if there is sufficient support to assume that samples are from the same distribution over multiple surveys. Specifically, for the four partnership distributions (displayed in Table S1), we compared three candidate assumptions with different combinations of shared parameters between datasets: the same parameters for all the three datasets (“All same”); different parameters for all the three datasets (“All different”); and the same parameters for Natsal-2 and 3 and different for Natsal-COVID (“Only 2020 different”). We used the Widely-applicable Bayesian Information Criterion (WBIC) (*34*) to compare models based on these assumptions. We considered a difference in WBIC of 2 or greater as positive support and 6 or greater as strong support (*35*). The results suggested that the same-sex partnership data reported by men was most likely derived from the same distribution throughout the three Natsal datasets and that the opposite-sex partnership data reported by men likely had different distributions between Natsal-2/3 and Natsal-COVID. The opposite-sex partnership reported by women was strongly suggested to have followed different distributions between the three surveys.

Assuming that the suggested deviation in the opposite-sex partnership distribution for both men and women reported in 2020 from the previous years may reflect the lockdown-associated behaviour changes, we used the all three Natsal datasets for MSM, Natsal-2 and 3 for HS men and Natsal-3 for HS women to estimate the partnership distributions in our main analysis. Results for alternative combinations of the datasets (pre-COVID and COVID wave 1 datasets) were also explored as part of our sensitivity analysis (*Sensitivity analysis 3*).

**Table S2.** Model selection to check for consistency between Natsal datasets.

| Category of partners | WBIC values for different assumptions |  |  |
| --- | --- | --- | --- |
|  | All same | All different | Only 2020 different |
| Same sex, reported by men | **1919.0 (best)** | 1932.2 ($\Delta$ 13.2) | 1933.3 ($\Delta$ 14.3) |
| Opposite sex, reported by men | 18873.5 ($\Delta$ 26.8) | 18858.9 ($\Delta$ 12.2) | **18846.7 (best)** |
| Same sex, reported by women | **790.3 (best)** | 797.3 ($\Delta$ 7.0) | 792.2 ($\Delta$ 1.9) |
| Opposite sex, reported by women | 17869.0 ($\Delta$ 32.3) | **17832.7 (best)** | 17877.5 ($\Delta$ 44.8) |

WBIC: Widely-applicable Bayesian Information Criterion. $\Delta$ : difference from the best model. The models with the best WBIC are highlighted in bold.

#### Maximum number of partners predicted by Pareto distributions fitted to empirical MSM sexual partnership data in previous studies

We simulated the Pareto-distributed number of sexual partners based on the estimates of exponent parameters reported in previous studies (*20, 21*) and compared the maximum number of partners reported in the datasets and that in the simulated samples. Parameter estimates in those studies are presented as an exponent parameter *γ*, which corresponds to *κ* + 1 in our parameterisation in Equation (6). For each of the dataset, a set of samples with a size equal to that of the original dataset was repeatedly drawn 1,000 times and the quantiles of the maximum number of partners among each sample set are compared with the original (Table S1). The Pareto distribution based on the parameter estimate for each of the dataset tended to produce substantially larger values for the maximum number of sexual partners among the simulated samples than observed. The observed maximum values in the three datasets (Natsal-2, Natsal-3 and GRASP) were all around the lower 2.5^th^ percentile of the Pareto-simulated maxima or below and smaller than the simulated medians by an order of magnitude or two. These results suggest that the Pareto distributions used to describe the sexual partnership distributions among MSM in the previous studies failed to capture the tail part of the empirical data and that a distribution with a more modest behaviour at the tail such as the Weibull distribution is expected to better characterise the observed sexual partnership distribution.

**Table S3.**
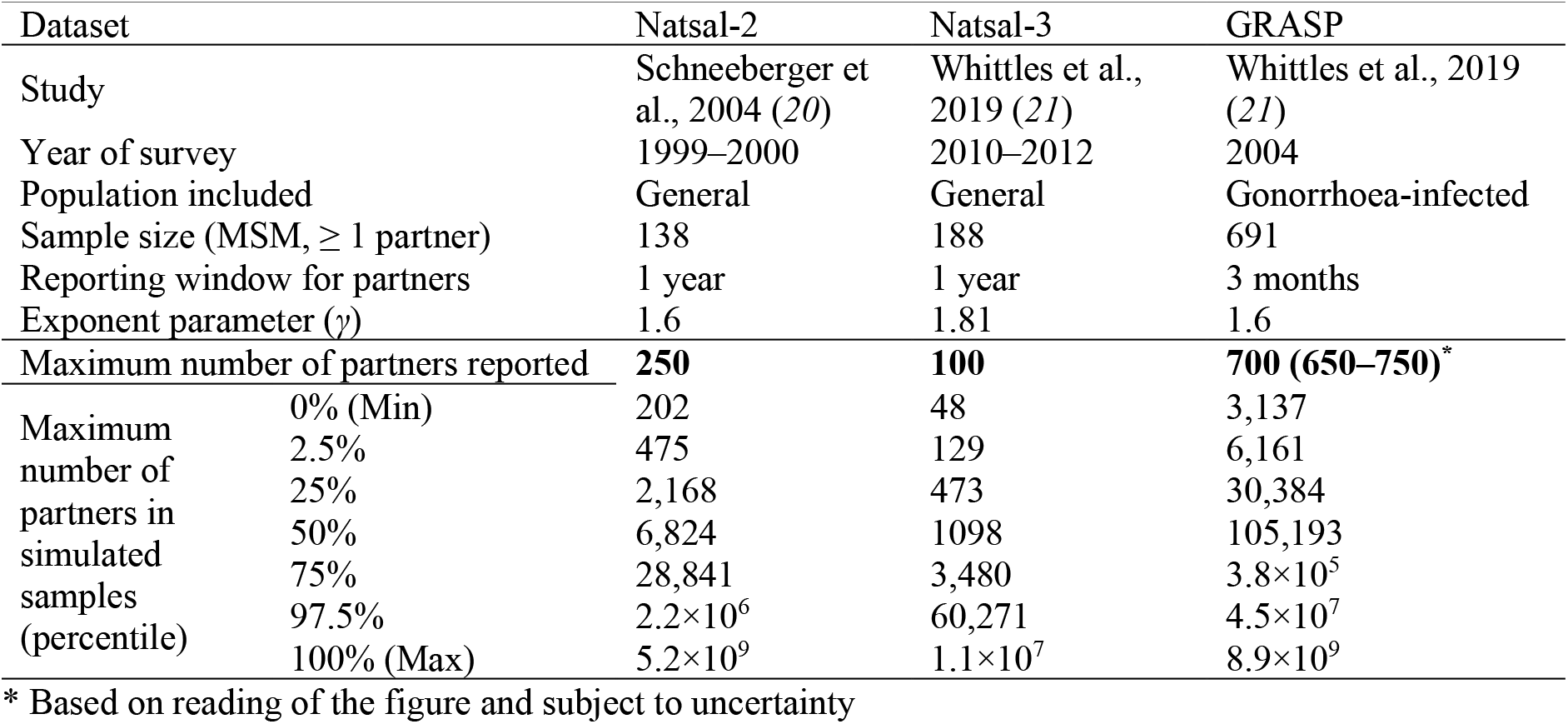
Maximum number of sexual partners among the MSM population in empirical and simulated datasets assuming a Pareto distribution.

| Dataset | Natsal-2 | Natsal-3 | GRASP |
| --- | --- | --- | --- |
| Study | Schneeberger et al., 2004 (20) | Whittles et al., 2019 (21) | Whittles et al., 2019 (21) |
| Year of survey | 1999–2000 | 2010–2012 | 2004 |
| Population included | General | General | Gonorrhoea-infected |
| Sample size (MSM, $\geq 1$ partner) | 138 | 188 | 691 |
| Reporting window for partners | 1 year | 1 year | 3 months |
| Exponent parameter ($\gamma$) | 1.6 | 1.81 | 1.6 |
| Maximum number of partners reported | **250** | **100** | **700 (650–750)\*** |
| Maximum number of partners in simulated samples (percentile) |  |  |  |
| 0% (Min) | 202 | 48 | 3,137 |
| 2.5% | 475 | 129 | 6,161 |
| 25% | 2,168 | 473 | 30,384 |
| 50% | 6,824 | 1098 | 105,193 |
| 75% | 28,841 | 3,480 | $3.8 \times 10^5$ |
| 97.5% | $2.2 \times 10^6$ | 60,271 | $4.5 \times 10^7$ |
| 100% (Max) | $5.2 \times 10^9$ | $1.1 \times 10^7$ | $8.9 \times 10^9$ |

\* Based on reading of the figure and subject to uncertainty

#### Sensitivity analysis 1: outbreak size in the UK

In the main analysis, we discussed the outbreak of monkeypox over sexual network in the global context, assuming the sexual partnership distributions in the UK are also representative of behaviours in other countries. Here, we shifted our focus to the outbreak within the UK, where the estimated sexual partnership distributions would most likely apply, by using the number of reported cases in the UK as of 31 May 2022 (190 cases). The results were almost identical to the main analysis (Table S3), suggesting that our conclusions would plausibly apply to the domestic outbreak in the UK (and other countries with a similar local outbreak size and sexual partnership distributions comparable to those in the UK).

**Table S3.** Likelihood of an outbreak over MSM or non-MSM sexual contact network given different numbers and profiles of introduction events, using UK outbreak size data.

| SAR | Likelihood of 190+ cases in MSM given 1 introduction event |  |  | Likelihood of 10000+ cases given multiple introductions |  |
| --- | --- | --- | --- | --- | --- |
|  | S-A event (MSM) | Non-S-A event (MSM) | Non-S-A event (Gen. pop.) | 728 S-A events in MSM | 1000 non-SA events in non-MSM |
| 5% | 10% | 0.26% | 0.007% | 100% | < 0.001% |
| 10% | 20% | 0.9% | 0.02% | 100% | < 0.001% |
| 20% | 32% | 2.6% | 0.05% | 100% | < 0.001% |
| 50% | 45% | 7.4% | 0.15% | 100% | < 0.001% |

SAR: secondary attack risk; MSM: men who have sex with men; S-A: Sexually-associated; Gen. pop.: general population.

#### Sensitivity analysis 2: Variations in Weibull parameters

We assessed the sensitivity of *R*_0_ over MSM and one-way HS reproduction numbers for HS men and women to variations in parameter estimates for the Weibull distribution fitted to the sexual partnership distributions (Fig. S2). The Pareto-approximated exponent *κ* was varied within the credible intervals in Table S1 and the shape parameter *α* was varied between 0.001–1.0 and transformed into the upper 1^st^ percentile of the number of partners (among those with non-zero partners) over 21 days.

**Fig. S2.**
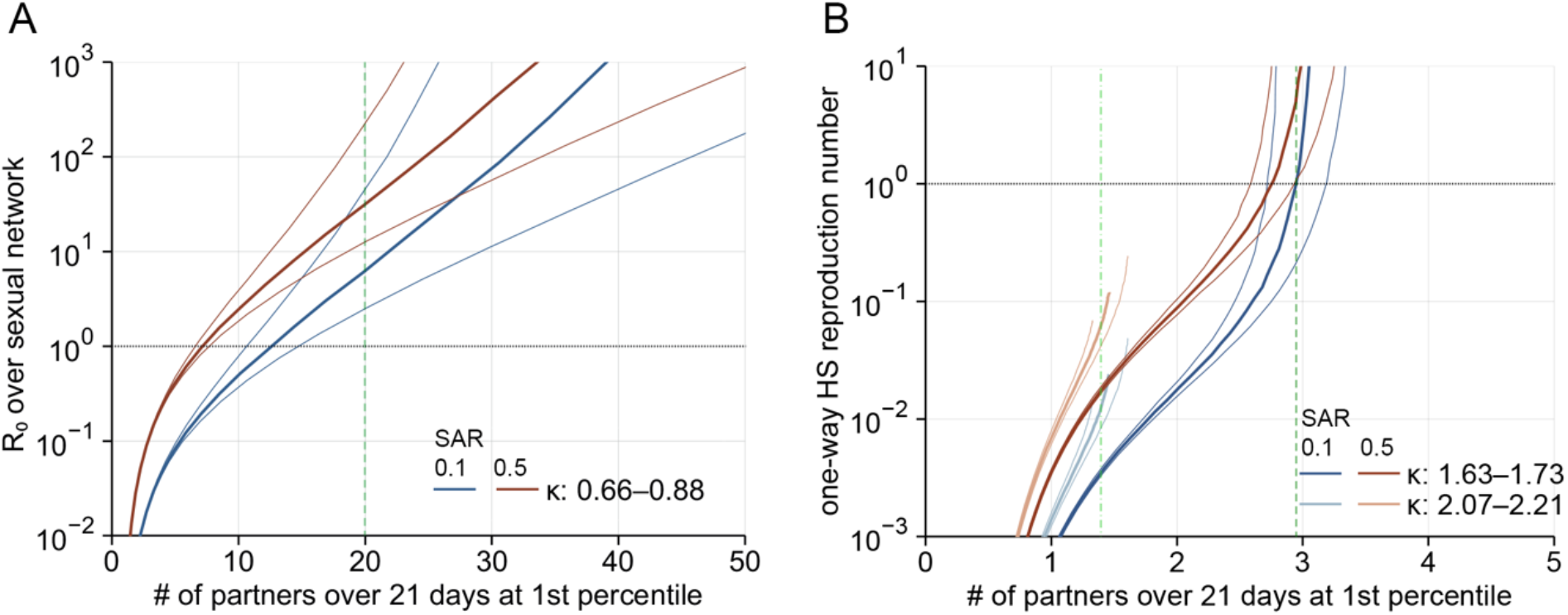
Sensitivity to variations in Weibull parameters for MSM and HS partnership distributions. (A) Projected *R*_0_ with the Weibull parameters varied within the 95% credible intervals for the sexual partnership distribution among MSM. The central thick lines correspond to the median estimates for *κ* and thinner lines the credible interval limits. The dotted horizontal lines denote the epidemic threshold (*R*_0_ = 1). Dashed green line indicates the upper 1^st^ percentile of the partnership distribution among those with non-zero partners. (B) Projected one-way HS reproduction numbers (*R*_FM_: darker lines; *R*_MF_: lighter lines) with the Weibull parameters varied within the credible intervals for the sexual partnership distributions among HS men and women. Dashed/dash-dotted green lines indicate the upper 1^st^ percentiles for HS men (dashed darker green) and women (dash-dotted lighter green). The open end of the curves corresponds to the lower limit of *α* = 0.001.

#### Sensitivity analysis 3: Infectious period and pre-COVID / COVID wave 1 datasets

We varied our assumption of 21 days for the infectious period of monkeypox as this estimate is based on the duration of illness from limited empirical data (*26*–*28*). The effective infectious period could also be shorter if individuals with symptoms refrain from having sexual contact. We used alternative assumptions of the infectious period (7, 14 and 28 days) and compared the *R*_0_ and the relative reduction required for control (Figs. S3A and S3B). The results suggest that the *R*_0_ for the non-MSM sexual contact network is below 1 for the entire range of SAR values if the infectious period is 7 or 14 days in this setting, while the *R*_0_ exceeds 1 with an SAR of ∼ 50% or higher if the infectious period is 28 days.

**Fig. S3.**
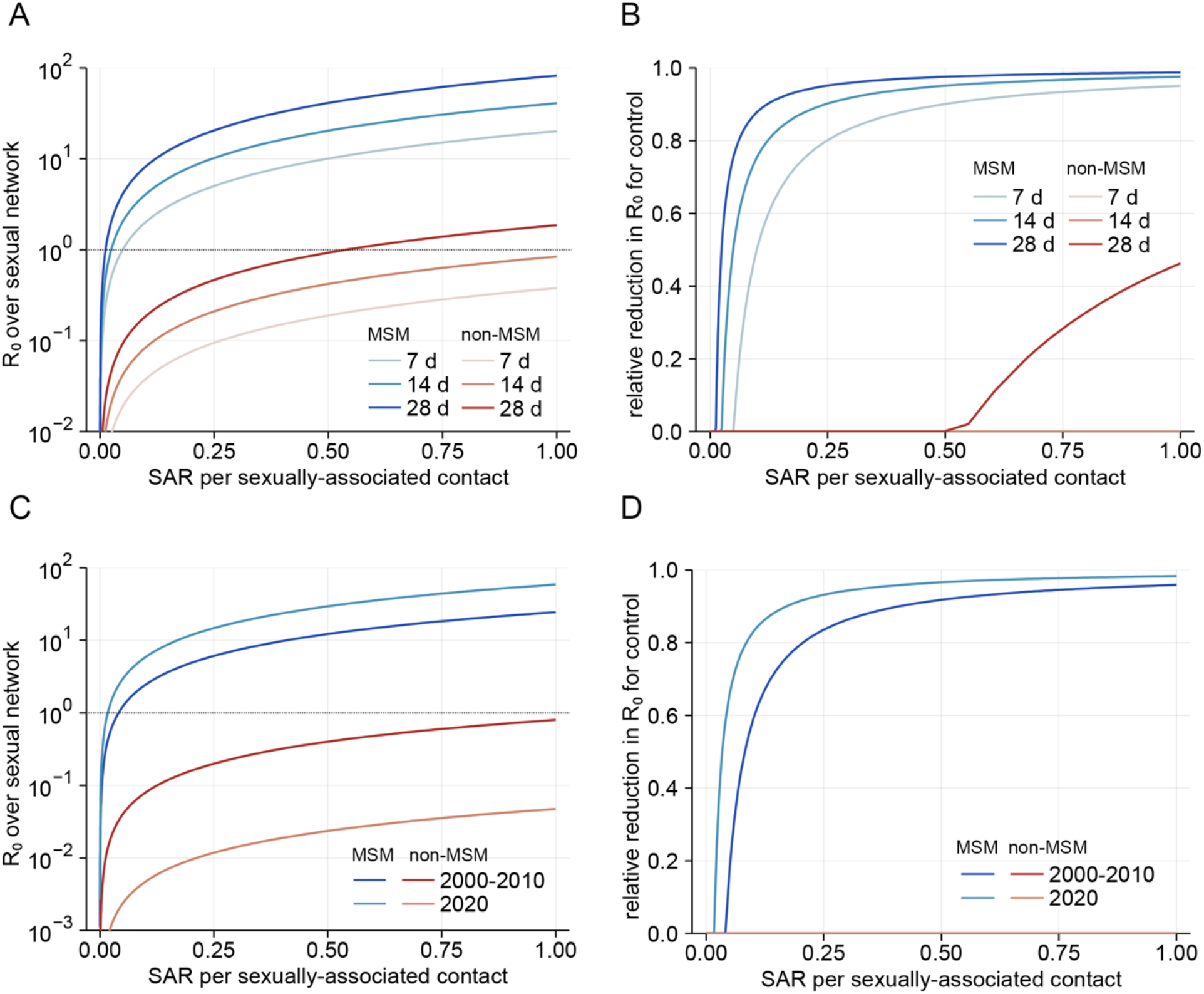
*R*_0_ of monkeypox over sexual contact networks and relative reduction required for control based on alternative infectious periods and datasets. (A) Projected *R*_0_ over the MSM and non-MSM sexual contact networks and (B) relative reduction in *R*_0_ required to bring the outbreak under control, assuming shorter and longer infectious periods (7 days, 14 days and 28 days) than the main analysis. (C) Projected *R*_0_ over the MSM sexual contact networks and (D) relative reduction in *R*_0_ required to bring the outbreak under control, based on alternative subsets of the Natsal data. The estimates from the pre-COVID datasets (Natsal-2/3, “2020-2010”) and the COVID wave 1 data (Natsal-COVID, “2020”) were used.

We also compared estimates obtained from alternative combinations of datasets, i.e. pre-COVID (Natsal-2 and 3, conducted in 1999–2000 and 2010–2012, respectively) and COVID wave 1 (Natsal-COVID conducted in summer 2020) datasets (Figs. S3C and S3D), to assess how having 3 months under lockdown prior to the Natsal-COVID survey might affect the estimated *R*_0_ of monkeypox via possible deviations in the sexual partnership distributions. The model comparison supported that the parameters for MSM be shared between the three datasets and the parameters for non-MSM (HS men and women) be separated between Natsal-2 and/or 3 and Natsal-COVID (Table S2). However, it is of note that not selecting the Natsal-COVID dataset to characterise the current sexual contact patterns among non-MSM (which we assumed are not under the effect of COVID-related changes) in the main analysis is only an assumption. The results of the sensitivity analysis suggested that using either the Natsal-2/3 or Natsal-COVID datasets instead of the three datasets combined for MSM does not lead to a substantial qualitative change. On the other hand, using the Natsal-COVID dataset for the non-MSM sexual partnership distribution caused an over 10-fold reduction in the estimated *R*_0_ over the non-MSM sexual network, which probably reflects the reduction in the tail-part of the partnership distribution among non-MSM during the lockdown.

